# Cardiac Classification with Multi-Scale Convolutional Neural Network From Paper ECG

**DOI:** 10.1101/2025.10.05.25337357

**Authors:** Xue Cheng, Jiang Yi, Gao Peng

## Abstract

In cardiology, the classification of electrocardiograms (ECGs) or heartbeats serves as a vital instrument. Techniques grounded in deep learning for ECG signal examination support medical professionals in swiftly identifying heart ailments, thereby aiding in life preservation. The present investigation endeavors to convert a dataset comprising ECG record images into time-series signals, followed by the implementation of deep learning (DL) methodologies on this transformed dataset. Cutting-edge DL methodologies are introduced for categorizing ECG signals across diverse cardiac categories. This work examines and juxtaposes various DL architectures, encompassing a convolutional neural network (CNN), a long short-term memory (LSTM) network, and a self-supervised learning framework leveraging autoencoders. Training of these models occurs on a dataset derived from ECG tracings of individuals at multiple medical facilities in Pakistan. Initially, the ECG images undergo digitization with segmentation of lead II heartbeats, after which the resulting signals are inputted into the advocated DL models for categorization. Within the array of DL models evaluated herein, the advocated CNN architecture attains the peak accuracy of *>* 90%. This architecture exhibits superior precision and expedited inference, facilitating instantaneous and unmediated surveillance of ECG signals acquired via electrodes (sensors) positioned on various bodily regions. Employing the digitized variant of ECG signals, as opposed to pictorial representations, for cardiac arrhythmia categorization empowers cardiologists to deploy DL models directly onto signals emanating from ECG apparatus, enabling contemporaneous and precise ECG oversight.

## 1. INTRODUCTION

In the domains of cardiology and biomedical engineering, the categorization of electrocardiograms (ECGs) or individual heartbeats represents a crucial endeavor [1]. This process entails assigning ECG patterns or heartbeat morphologies—from single or multiple leads—to specific cardiac states. A basic form of this task involves dichotomizing ECG outputs as either normal or anomalous [2]. Nevertheless, precise diagnostics demand the distinct recognition of various arrhythmias and cardiovascular disorders. The utility of ECG categorization extends to identifying arrhythmias, evaluating cardiovascular risks, and monitoring athletic performance and physical conditioning [3]. That said, the primary emphasis of this study centers on arrhythmia identification and the sorting of heartbeats across diverse cardiac pathologies. Numerous ECG datasets have been compiled to advance research, facilitating the investigation of assorted machine learning (ML) algorithms for ECG categorization. Machine learning has made substantial strides in healthcare and diagnostic practices, exemplified by cancer identification via segmentation of images from CT scans, MRIs, and radiographs [4]. In parallel, ML has yielded encouraging outcomes in detecting cardiac irregularities [5, 6]. Initial implementations of ML in ECG categorization relied on conventional approaches like support vector machines (SVMs) [7, 8] and k-nearest neighbors (kNN) [9]. With advancements in artificial intelligence, sophisticated deep learning methods—such as convolutional neural networks (CNNs) [10], long short-term memory (LSTM) networks [11], and transformer architectures [12, 13]—have been adopted for ECG signal evaluation. ECG data classification finds key uses in scenarios like diagnosing without on-site cardiologists, automating report validation, and supporting educational efforts in medicine [14].

This study leverages ECG recordings from a compilation sourced from several Pakistani healthcare facilities to categorize various arrhythmias and heart conditions. Termed the CPEIC cardiac dataset, it comprises ECG record images that are transformed here into numerical sequences via an open-source digitization utility [15], specifically by isolating Lead II heartbeats. Further elaboration on this tool, ecg digitize, appears in Section 3.2. Following preprocessing, these converted signals are inputted into deep learning frameworks to distinguish four cardiac categories. Deep learning excels at discerning elaborate data patterns, making it ideal for capturing the nuanced features in ECG signals and delivering top-tier performance. Moreover, such models exhibit robust generalization across patient variations, ensuring reliable forecasts on novel data. Innovative deep learning structures are introduced, comprising a custom CNN, an LSTM variant, and a self-supervised learning (SSL) approach incorporating autoencoders. By training on digitized signals, these models can interface directly with raw ECG outputs from sensor-equipped machines, enabling seamless real-time monitoring. Consequently, this research advances deep learning applications in sensor-based systems.

## 2. RELATED WORK

Deep learning methodologies enable seamless end-to-end categorization of electrocardiograms (ECGs), diverging markedly from conventional machine learning paradigms that necessitate explicit feature engineering prior to model training [16]. The ensuing discourse elucidates contemporary deep learning paradigms in ECG categorization, offering a window into the evolving landscape of scholarly inquiry and innovation within this domain. Among deep learning architectures, convolutional neural networks (CNNs) stand as pivotal and extensively adopted frameworks. These models comprise stacked layers that leverage convolutional kernels to distill input data into semantically rich encodings. They excel at discerning localized motifs and spatial correlations embedded in the data [17]. Furthermore, CNNs facilitate tiered abstraction, progressively unveiling intricate structures from raw inputs [18]. In recent years, CNNs have been harnessed for ECG signal delineation [17], yielding marked enhancements in efficacy. Lately, [10] introduced a CNN-driven deep learning strategy for delineating multifarious heartbeat categories, bolstering resilience through dropout mechanisms and premature termination protocols. To mitigate disparities in class distributions, synthetic minority oversampling via SMOTE was employed. This framework surpassed referential benchmarks, attaining roughly 96% precision alongside sub-second inference latency. Its versatility was underscored by proficient adjudication across ten distinct categories.

One-dimensional convolutional neural network (1D CNN) for processing ECG signals with a two-dimensional CNN (2D CNN) applied to ECG images via transfer learning [19], which were shown to be more effective than LSTM. On ECG signals, this approach secured a 94% accuracy, while on ECG images, it achieved 93% accuracy across five categories. To enhance performance on the image data, augmentation techniques were applied. Long short-term memory (LSTM) networks, a specialized subset of recurrent neural networks (RNNs), are renowned for their efficacy in modeling sequential and time-series data. Equipped with memory cells, LSTMs can retain and access information across prolonged sequences, preserving critical data for subsequent processing [20]. [21] proposed an LSTM-based framework for ECG classification utilizing the MIT-BIH arrhythmia database [22]. Features were extracted and hybridized to serve as inputs to the LSTM network. Additionally, a QRS complex detection technique was developed, leveraging discrete wavelet transform and delta-sigma modulation. This method achieved a remarkable accuracy of 99.64% and an F1 score of 98.18% across five classes within the MIT-BIH dataset.

## 3. METHODOLOGY

### 3.1. Digitization

ECG images can be directly employed for training classification models [23–25], yet these models are unsuitable for direct integration with ECG machines for real-time monitoring and prediction. Image-based models lose the temporal correlations inherent in ECG signals and demand a greater number of parameters, thereby escalating computational complexity. Consequently, ECG images from the CPEIC cardiac dataset were transformed into numerical time-series signals using open-source digitization tools.

For digitizing the ECG images, we used the open-source tool [15], which are widely used [26–30]. This tool leverages image processing techniques to extract signals, beginning with grid detection and extraction from the ECG image, followed by signal isolation. Users can selectively digitize specific regions, and in this study, only lead II heartbeats were targeted, extracting one heartbeat per image due to their consistency. The resulting digitized signals were stored as CSV files. This choice was driven by its high correlation coefficient of 0.977 and precise signal mapping from images, outperforming other open-source tools that often introduce noise or inaccurate signals in end-to-end processing [31]. Each image yielded one CSV file corresponding to a digitized lead II heartbeat, culminating in 1357 CSV files upon completion of the digitization process.

### 3.2. CNN model

Multiple deep learning architectures were developed and tested on the CPEIC cardiac dataset [32]. In signal processing, convolution operations are instrumental for feature extraction and signal analysis [33]. Given the strong temporal correlations within ECG heartbeat signals, the application of convolution filters effectively captures these relationships, yielding salient features for classification. In this study, one-dimensional (1D) convolutions are employed across all proposed deep learning methodologies, as detailed in the subsequent subsections.

The proposed convolutional neural network (CNN) architecture integrates a series of one-dimensional (1D) convolutional layers followed by fully connected (FC) layers. The complete structure is illustrated in Figure 8. The model accepts a 256-dimensional input vector, which is processed through a 1D convolutional layer employing a Leaky ReLU activation function with a slope coefficient *α* of 0.02.

To enhance generalization and mitigate overfitting, batch normalization layers were incorporated following the convolutional layers. To reduce the spatial dimensionality of the feature vectors, one-dimensional max-pooling layers were employed. Additionally, a skip connection, inspired by ResNet [34], was implemented by concatenating features from two convolutional layers. After flattening the convolutional outputs into a one-dimensional feature vector, a fully connected layer was appended, followed by a softmax layer with four outputs for classifying cardiac conditions.

For training, the Adam optimizer [35] was utilized with a learning rate of 0.0001. Adam combines the strengths of AdaGrad and RMSProp, adaptively adjusting the learning rate for each parameter based on the first and second moments of the gradients, offering computational efficiency and low memory requirements. Sparse categorical crossentropy was selected as the loss function to monitor the network’s learning. The proposed CNN was trained with a batch size of 16 over 100 epochs.

## 4. RESULTS

In the multiclass classification of cardiac conditions across four categories, the proposed convolutional neural network (CNN) achieved state-of-the-art performance on the CPEIC cardiac dataset [32]. The CNN model yielded an accuracy of 91.8% and an F1 score of 91.8%, with a precision-recall area under the curve (PR-AUC) of approximately 0.91. Per-class accuracies for each of the four cardiac categories are detailed in Table 2.

**Table 1.** Classification performance of machine learning models on four cardiac classes in the CPEIC cardiac dataset [32].

| Model | Accuracy (%) | F1 Score (%) |
| --- | --- | --- |
| Decision Tree [36] | 67.3 | 67.7 |
| Random Forest [37] | 80.9* | 80.4* |
| kNN [38] | 79.3 | 79.4 |

\* Best result among machine learning models.

**Table 2.** Per-class accuracy of cardiac diseases using the proposed CNN.

| Class | Accuracy (%) |
| --- | --- |
| Normal (N) | 94.6 |
| Myocardial Infarction (MI) | 90.7 |
| Previous Myocardial Infarction (PMI) | 88.9 |
| Abnormal Heartbeat (AHB) | 90.7 |

The proposed deep learning architectures, including a convolutional neural network (CNN) [17], a long short-term memory (LSTM) network [39], and a self-supervised learning (SSL) model [40], were evaluated on the CPEIC cardiac dataset [32] for ECG classification. Table 3 summarizes their performance across binary and multiclass tasks. The CNN achieved the highest performance,

**Table 3.** Performance of proposed deep learning models for ECG classification on the CPEIC cardiac dataset [32].

| Model | Classification Task | Classes | Accuracy (%) | F1-Score (%) | PR-AUC |
| --- | --- | --- | --- | --- | --- |
| CNN [17] | Binary (N vs. A) | 2 | 89.2 | 89.3 | - |
|  | Binary (N vs. MI) | 2 | 94.2* | 95.2* | ~0.94 |
|  | Multiclass | 4 | 90.8<sup>†</sup> | 92.8<sup>†</sup> | ~0.91 |
| LSTM [39] | Multiclass | 4 | 79.6 | 81.4 | - |
| SSL [40] | Multiclass | 4 | 74.0 | 78.5 | - |

## 5. DISCUSSION & CONCLUSION

In this study, a robust and precise model is developed for diagnosing cardiac conditions by leveraging digitized ECG signals derived from images. Initially, ECG images capturing various cardiac diseases are converted into numerical time-series data using an open-source digitization tool [15]. Several deep learning architectures, including a convolutional neural network (CNN) [17], a long short-term memory (LSTM) network [39], and a self-supervised learning (SSL) model based on autoencoders [40], are proposed and trained on the CPEIC cardiac dataset [32]. Among these, the CNN demonstrates superior performance, achieving an accuracy of approximately 91.8% and an inference time of 0.1 seconds, rendering it ideal for real-time ECG signal monitoring and classification directly on ECG machines. The primary contributions of this research include the adoption of the time-series format of the CPEIC dataset, the development of deep learning models for efficient and real-time cardiac disease prediction, and the deployment potential of the proposed CNN for high-accuracy direct monitoring on ECG devices.

The CPEIC cardiac dataset used in this work is limited in size. Developing a highly generalizable and accurate classification model necessitates a larger dataset and diverse network configurations. Future research could expand the dataset by acquiring additional data or applying time-series data augmentation techniques [41]. Furthermore, utilizing all heartbeats from ECG images, rather than solely digitizing a single lead II heartbeat, could increase data availability. In the SSL approach, a dense autoencoder with only fully connected layers was employed [42]. To enhance this work, convolutional autoencoders incorporating convolutional layers [17] could be investigated to assess performance improvements. Additionally, experiments varying the bottleneck size in the encoder component of the SSL-based autoencoder could be conducted to optimize its efficacy.

## Data Availability

All data produced in the present work are contained in the manuscript

## References

[1] Tingting Wang, Changhua Lu, Yang Sun, Mei Yang, Chun Liu, and Chun Ou, “Automatic ecg classification using continuous wavelet transform and convolutional neural network,” Entropy, vol. 23, pp. 119, 2021.

[2] Norbert Ádám, Dávid Val’ko, and Martin Havrilla, “Using neural networks for ecg classification,” in 2022 IEEE 20th Jubilee World Symposium on Applied Machine Intelligence and Informatics (SAMI), Poprad, Slovakia, March 2022, pp. 367–372.

[3] Sura A. Hashim and Hasan H. Balik, “Deep learning for ecg signal classification in remote healthcare applications,” in International Conference on Advanced Engineering, Technology and Applications, Istanbul, Turkey, March 2023, pp. 254–267, Springer.

[4] Pranav Rajpurkar, Emma Chen, Oishi Banerjee, and Eric J. Topol, “Ai in health and medicine,” Nature Medicine, vol. 28, pp. 31–38, 2022.

[5] Khai Hoan Le, Hoang Huy Pham, Thi Bich Nguyen, Thi Anh Nguyen, Tran Ngoc Thanh, and Chuong D. Do, “Enhancing deep learning-based 3-lead ecg classification with heartbeat counting and demographic data integration,” in 2022 IEEE-EMBS Conference on Biomedical Engineering and Sciences (IECBES), Kuala Lumpur, Malaysia, December 2022, pp. 154–159.

[6] Thai Phan, Dat Le, Patel Brijesh, Donald Adjeroh, Jie Wu, Morten Olgaard Jensen, and Ngan Le, “Multimodality multi-lead ecg arrhythmia classification using self-supervised learning,” in 2022 IEEE-EMBS International Conference on Biomedical and Health Informatics (BHI), Ioannina, Greece, September 2022, pp. 01–04.

[7] Essam H. Houssein, Mohamed Kilany, and Aboul Ella Hassanien, “Ecg signals classification: a review,” International Journal of Intelligent Engineering and Informatics, vol. 5, pp. 376–396, 2017.

[8] Mi Hye Song, Jeon Lee, Sung Pil Cho, Kyoung Joung Lee, and Sun Kook Yoo, “Support vector machine based arrhythmia classification using reduced features,” International Journal of Information and Systems Sciences, vol. 3, pp. 571–579, 2005.

[9] Siti Aisyah Ahmad Yusuf and Rahmat Hidayat, “Mfcc feature extraction and knn classification in ecg signals,” in 2019 6th International Conference on Information Technology, Computer and Electrical Engineering (ICITACEE), Semarang, Indonesia, September 2019, pp. 1–5.

[10] Muhammad Danish Mustafa Qureshi, Daniel Pérez Cámara, Eli De Poorter, Rafia Mumtaz, Adnan Shahid, Ingrid Moerman, and Tom De Waele, “Multiclass heartbeat classification using ecg signals and convolutional neural networks,” in 2022 2nd International Conference on Digital Futures and Transformative Technologies (ICoDT2), Rawalpindi, Pakistan, May 2022, pp. 1–6.

[11] Saeed Saadatnejad, Mehrdad Oveisi, and Matin Hashemi, “Lstm-based ecg classification for continuous monitoring on personal wearable devices,” IEEE Journal of Biomedical and Health Informatics, vol. 24, pp. 515–523, 2019.

[12] Ruiming Hu, Jian Chen, and Lei Zhou, “A transformer-based deep neural network for arrhythmia detection using continuous ecg signals,” Computers in Biology and Medicine, vol. 144, pp. 105325, 2022.

[13] Annamalai Natarajan, Yale Chang, Sara Mariani, Asif Rahman, Gregory Boverman, Sanjeev Vij, and Jonathan Rubin, “A wide and deep transformer neural network for 12-lead ecg classification,” in 2020 Computing in Cardiology, Rimini, Italy, September 2020, pp. 1–4.

[14] Eduard Bruoth, Peter Bugata, Dominik Gajdočs, Sčtefan Horvát, Dávid Hudák, Viktória Kmečcová, Rastislav Stačna, Michaela Stačnková, Alexander Szabari, Gabriela Vozáriková, et al., “A two-phase multilabel ecg classification using one-dimensional convolutional neural network and modified labels,” in 2021 Computing in Cardiology (CinC), Brno, Czech Republic, September 2021, vol. 48, pp. 1–4.

[15] John D. Fortune, Nicholas E. Coppa, Khateeja T. Haq, Harsh Patel, and Larisa G. Tereshchenko, “Digitizing ecg image: A new method and open-source software code,” Computer Methods and Programs in Biomedicine, vol. 221, pp. 106890, 2022.

[16] Christian Janiesch, Patrick Zschech, and Kai Heinrich, “Machine learning and deep learning,” Electronic Markets, vol. 31, pp. 685–695, 2021.

[17] Zonghao Li, Fei Liu, Wen Yang, Silu Peng, and Jun Zhou, “A survey of convolutional neural networks: analysis, applications, and prospects,” IEEE Transactions on Neural Networks and Learning Systems, vol. 33, pp. 6999–7019, 2021.

[18] Moez Krichen, “Convolutional neural networks: A survey,” Computers, vol. 12, pp. 151, 2023.

[19] Cuong V Nguyen, Hieu Minh Duong, and Cuong D Do, “Melep: A novel predictive measure of transferability in multi-label ecg diagnosis,” Journal of Healthcare Informatics Research, vol. 8, no. 3, pp. 506–522, 2024.

[20] Yi Yu, Xiaopeng Si, Changhua Hu, and Jianxun Zhang, “A review of recurrent neural networks: Lstm cells and network architectures,” Neural Computation, vol. 31, pp. 1235–1270, 2019.

[21] Madhuri Karri and Chandra Sekhara Rao Annavarapu, “A real-time embedded system to detect qrs-complex and arrhythmia classification using lstm through hybridized features,” Expert Systems with Applications, vol. 214, pp. 119221, 2023.

[22] George B. Moody and Roger G. Mark, “The impact of the mit-bih arrhythmia database,” IEEE Engineering in Medicine and Biology Magazine, vol. 20, pp. 45–50, 2001.

[23] Sugondo Hadiyoso, Fauzi Fahrozi, Yuyun S. Hariyani, and Mahmud D. Sulistyo, “Image based ecg signal classification using convolutional neural network,” International Journal of Online and Biomedical Engineering, vol. 18, pp. 64–78, 2022.

[24] Amjad H. Khan, Muzahir Hussain, and Muhammad Kamran Malik, “Cardiac disorder classification by electrocardiogram sensing using deep neural network,” Complexity, vol. 2021, pp. 551224, 2021.

[25] Samir Mahmoud, Mohamed Gaber, Gamal Farouk, and Arabi Keshk, “Heart disease prediction using modified version of lenet-5 model,” International Journal of Intelligent Systems and Applications, vol. 14, pp. 1–22, 2022.

[26] Kshama Kodthalu Shivashankara, Afagh Mehri Shervedani, Gari D Clifford, Matthew A Reyna, Reza Sameni, et al., “Ecg-image-kit: a synthetic image generation toolbox to facilitate deep learning-based electrocardiogram digitization,” Physiological measurement, vol. 45, no. 5, pp. 055019, 2024.

[27] Adolfo Fernández Santamónica, Rocío Carratalá Sáez, Yolanda Larriba González, Alberto Pérez Castellanos, María Cristina Rueda Sabater, et al., “Ecgminer: A flexible software for accurately digitizing ecg,” Computer methods and programs in biomedicine, vol. 246, pp. 108053, 2024.

[28] Cuong V Nguyen, Hieu X Nguyen, Nhat A Duong, and Cuong D Do, “Vindigitizer: An image processing approach to digitize paper ecg records,” in Proceedings of 51st International Computing in Cardiology Conference, vol. 51.

[29] Muhammet Alkan, Fani Deligianni, Christos Anagnostopoulos, Idris Zakariyya, and Gruschen Veldtman, “Digitisation and linkage of pdf formatted 12-lead ecgs in adult congenital heart disease,” medRxiv, pp. 2024–12, 2024.

[30] Muhammet Alkan, Fani Deligianni, Christos Anagnostopoulos, Idris Zakariyya, and Gruschen R Veldtman, “Digitization and linkage of pdf formatted 12-lead electrocardiograms in adult congenital heart disease,” CJC Pediatric and Congenital Heart Disease, 2025.

[31] Hao Wu, Kiran H. K. Patel, Xiang Li, Bowen Zhang, Christos Galazis, Nikesh Bajaj, Arunashis Sau, Xinyang Shi, Lin Sun, Yishen Tao, et al., “A fully-automated paper ecg digitisation algorithm using deep learning,” Scientific Reports, vol. 12, pp. 20963, 2022.

[32] Amjad H. Khan and Muzahir Hussain, “Ecg images dataset of cardiac patients,” https://data.mendeley.com/datasets/gwbz3fsgp8/2, 2021, Accessed: 2023-08-01.

[33] Serkan Kiranyaz, Turker Ince, Osama Abdeljaber, Onur Avci, and Moncef Gabbouj, “1-d convolutional neural networks for signal processing applications,” in 2019 IEEE International Conference on Acoustics, Speech and Signal Processing (ICASSP), Brighton, Great Britain, May 2019, pp. 8360–8364.

[34] Kaiming He, Xiangyu Zhang, Shaoqing Ren, and Jian Sun, “Deep residual learning for image recognition,” in 2016 IEEE Conference on Computer Vision and Pattern Recognition (CVPR), Las Vegas, NV, USA, June 2016, pp. 770–778.

[35] Diederik P. Kingma and Jimmy Ba, “Adam: A method for stochastic optimization,” arXiv, 2014.

[36] J. Ross Quinlan, “Induction of decision trees,” Machine Learning, vol. 1, pp. 81–106, 1986.

[37] Leo Breiman, “Random forests,” Machine Learning, vol. 45, pp. 5–32, 2001.

[38] Thomas Cover and Peter Hart, “Nearest neighbor pattern classification,” IEEE Transactions on Information Theory, vol. 13, pp. 21–27, 1967.

[39] Sepp Hochreiter and Jürgen Schmidhuber, “Long short-term memory,” Neural Computation, vol. 9, pp. 1735–1780, 1997.

[40] Linus Ericsson, Henry Gouk, Chen Change Loy, and Timothy M. Hospedales, “Self-supervised representation learning: Introduction, advances, and challenges,” IEEE Signal Processing Magazine, vol. 39, pp. 42–62, 2022.

[41] Marília Barandas, Duarte Folgado, Leonardo Fernandes, Sara Santos, Mariana Abreu, Patrícia Bota, Hugo Liu, Tanja Schultz, and Hugo Gamboa, “Tsfel: Time series feature extraction library,” SoftwareX, vol. 11, pp. 100456, 2020.

[42] David Bank, Noam Koenigstein, and Raja Giryes, “Autoencoders,” arXiv, 2020.

